# The Lived Experience of Infertility in Asian Americans: A Qualitative Study

**DOI:** 10.1101/2024.09.21.24314139

**Authors:** Michelle Vu, Anh-Tho Antoinette Nguyen, Jinsol Hyun, Timothy Dye, Snigdha Alur-Gupta

## Abstract

**Objective:** To identify psychosocial and cultural factors among Asian American patients diagnosed with infertility that may contribute to their disparate fertility outcomes.

**Design:** A cross-sectional qualitative interview study was conducted with women ≥18-years-old who identified as Asian American and had a diagnosis of infertility. Semi-structured interviews were performed until thematic saturation was reached. The interviews were transcribed and coded. A codebook was developed to capture the grounded perspectives of participants. The web application Dedoose was utilized to conduct content analysis, identifying common themes and patterns.

**Results:** Asian Americans voice the importance of having children, emphasizing the cultural expectation to procreate. They reported that infertility caused immense mental and emotional distress, resulting in feelings of disappointment and grief. Participants noted that it is not common in their culture to talk about infertility-related issues. Religion and naturopathic remedies played a large role in helping participants adjust to their infertility journeys. Participants stated that the infertility evaluation was complicated and one particular area of frustration was the lack of individually centered treatment. Interestingly, participants expressed desire to help other Asian women who are also struggling with infertility.

**Conclusion:** Infertility is a difficult and complex experience for Asian American women. Parenthood is a strong expectation for people of Asian descent and results in emotional burden when complicated by infertility. Fertility concerns are considered taboo to openly discuss, which can cause additional feelings of isolation. Healthcare providers should work to address the culturally induced shame associated with infertility and provide more individualized care.

## Introduction

Infertility is a widespread disease affecting over 13% of women in the United States (U.S.).[1] In recent years, there has been a large increase in publications reporting racial and ethnic disparities in reproductive medicine.[2] Asian American infertility, however, is a particularly understudied area of research. “Asian” as a racial category encompasses a large and diverse contingent of people, including those with origins in East, Southeast, and South Asia.[3] According to the 2020 U.S. Census Bureau population estimate, there are over 24 million Asian Americans, accounting for over 7.2% of the nation’s population.[4] Despite the millions of Asian Americans seeking healthcare in the U.S., Asian Americans are underrepresented in health research, and are especially underrepresented in the infertility literature.[5, 6]

A few studies published on Asian American infertility reported that Asian American patients who decide to pursue fertility treatment have poorer outcomes for both intrauterine insemination (IUI) and in vitro fertilization (IVF) [7, 8] when compared with their white counterparts.[9, 10] In a study of the Society for Assisted Reproductive Technology (SART) database from 1999 to 2000 combined with data from the University of California, San Francisco reproductive health clinic from 2001 to 2003, Asian American women had decreased clinical pregnancy rates compared to white women. Multivariate analysis of the study data demonstrated that Asian ethnicity was an independent predictor of poor outcomes.[11] Additionally, Asian American patients are more likely to wait longer to seek infertility evaluation, presenting to fertility clinics at later ages.[7, 11] A retrospective cohort study of patients undergoing IUI at the University of California, San Francisco from 2002 to 2006 found that a greater proportion of Asian American patients (43.9%) presented for treatment after >2 years of infertility compared to white patients (24.6%) (p<0.0001).[8]

Many suspected sociocultural reasons could potentially explain the disparities in Asian American infertility and treatment outcomes. In qualitative study of 13 Chinese women residing in Hong Kong, the women reported feeling pressured to bear children and were frequently blamed for their inability to conceive.[12] Infertility is also associated with social stigma, and Asian women report a strong sense of shame associated with the diagnosis, resulting in psychologic distress.[6, 13] Asian Americans are also less likely to report strong physician-patient relationships. Ngo-Metzger et al. conducted a study administering the 2001 Health Care Quality Survey to both Asian Americans and non-Hispanic white patients. Asian Americans were more likely to report that their physicians did not understand their background and values, and their doctors did not listen or involve them in healthcare decisions as much as they wanted.[14]

In a literature review evaluating infertility among Asian Americans within the United States, only 12 studies were found in the past 15 years, and there are no published qualitative studies. We aimed to conduct interviews with Asian American patients facing infertility to assess the psychosocial and cultural factors that may explain the disparate fertility outcomes. A qualitative approach helps bridge a gap in the literature and identifies the experiences, attitudes, and beliefs of Asian American patients that might not be validly captured numerically.

## Materials and Methods

We performed an ethnographic qualitative interview study with the goal of allowing participants to explain how, why, or what they were thinking, feeling, and experiencing during their infertility journeys. Potential subjects were identified through an electronic medical record query at a large, academic medical institution. The inclusion criteria for the study included women 18 years and older, self-identified as Asian, with a diagnosis of infertility (ICD-10 code N97.9) and evaluated at one of the academic center’s resident obstetrics/gynecology clinics and the reproductive endocrinology and infertility (REI) clinic from 2016-2021. The exclusion criteria for the study included women who self-identified with another race or ethnicity, including patients of mixed race, and women who did not understand or speak English.

A qualitative discussion guide was created by the entire research team to help interview patients (Figure 1). The guide included questions about family building goals, challenges to fertility care, and experiences with the healthcare received. The questions were based on two previous qualitative studies with Asian patients experiencing infertility conducted in Kowloon, Hong Kong and Tianjin, China.[12, 18] 2 pilot interviews were conducted to ensure comprehensibility of the questions. 3 members of the research team conducted the interviews, including 1 obstetrics/gynecology resident (MV) and 2 medical students (AN, JH), all of whom identify as Asian American females. Subjects were called at the primary phone number listed in their charts. We thoroughly explained to subjects the purpose and goals of our study and they were asked if they were interested in participating. If yes, a consent form was read to the patient and a verbal consent was obtained and documented in RedCap.

**Figure 1.**
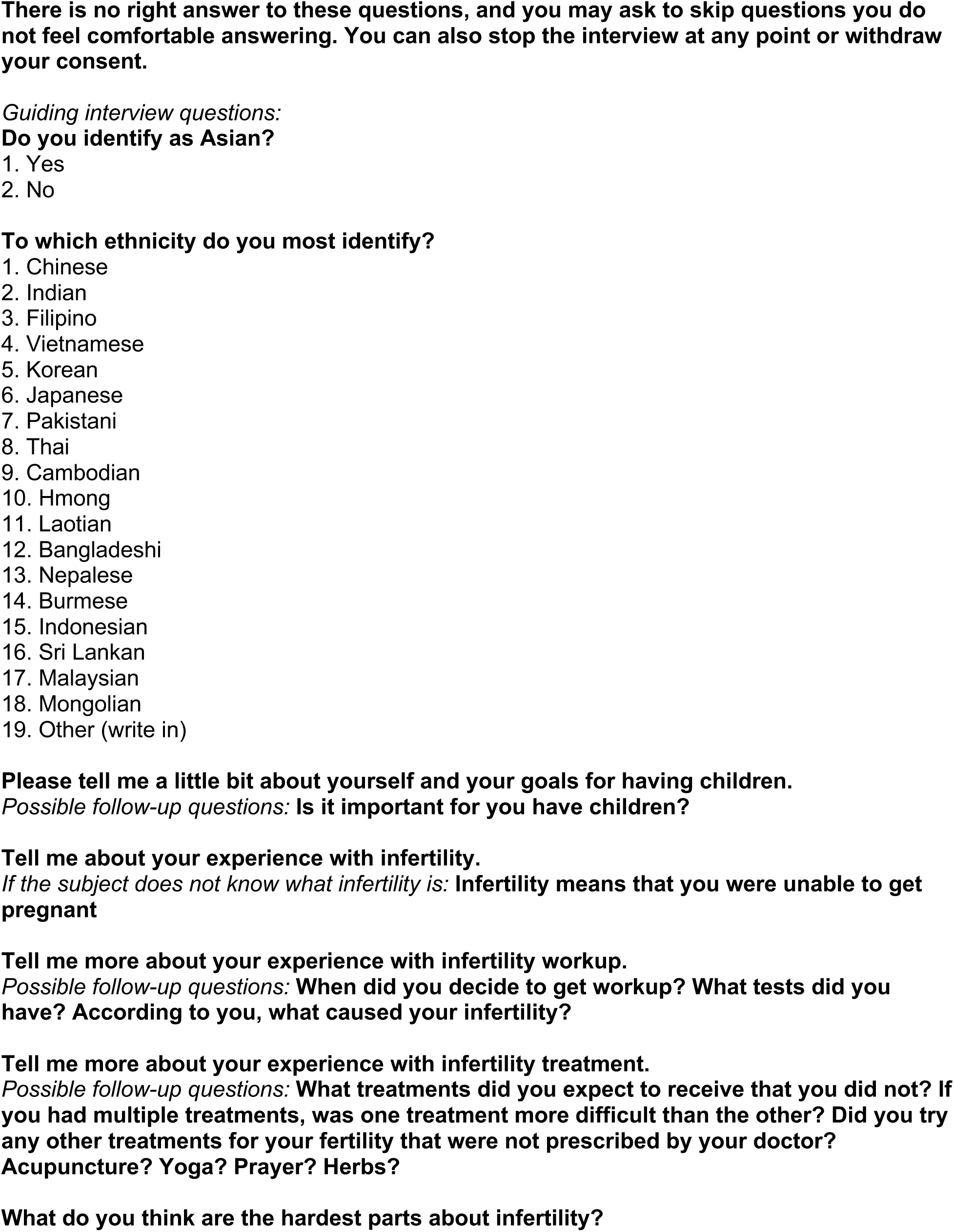
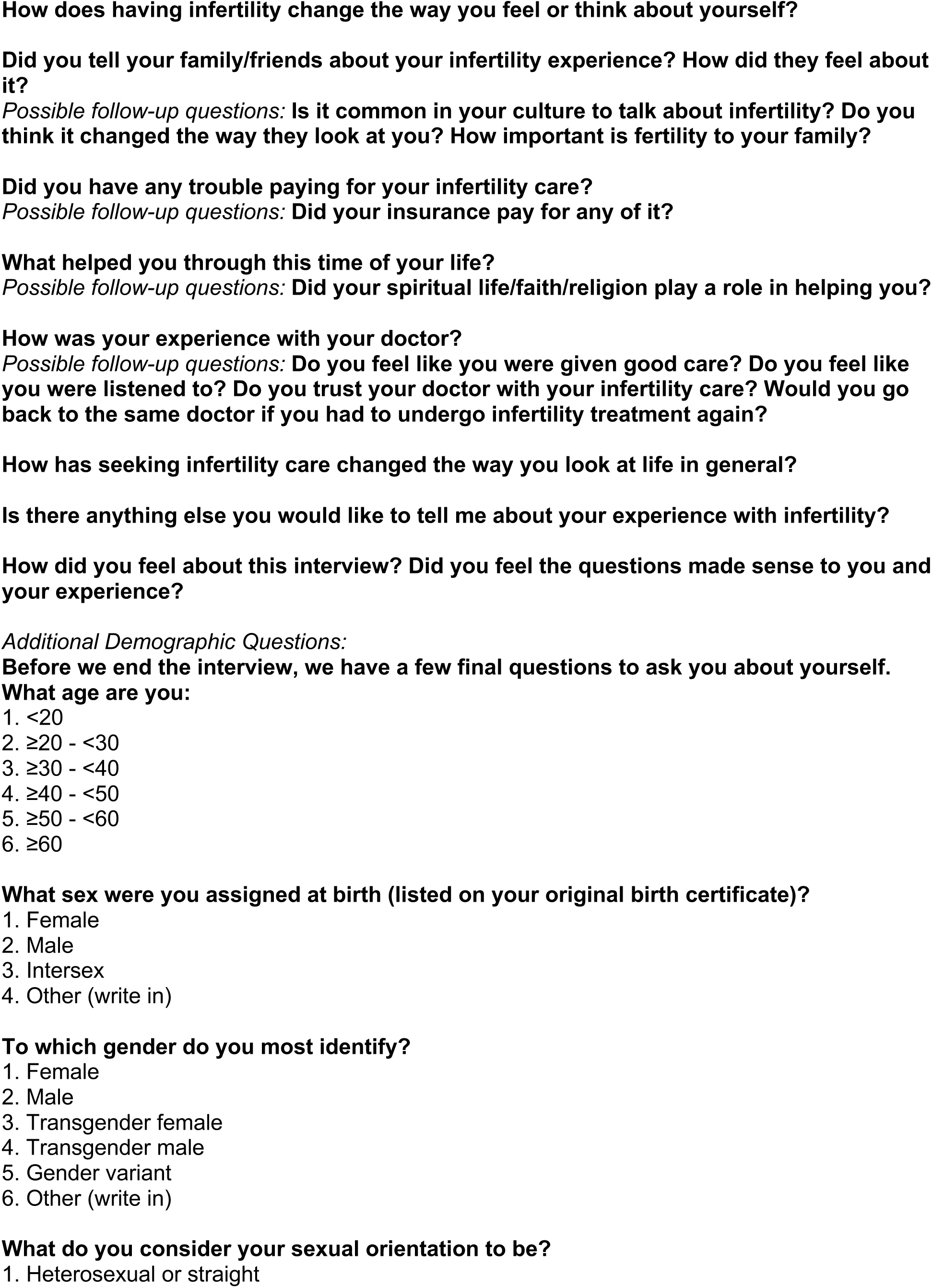
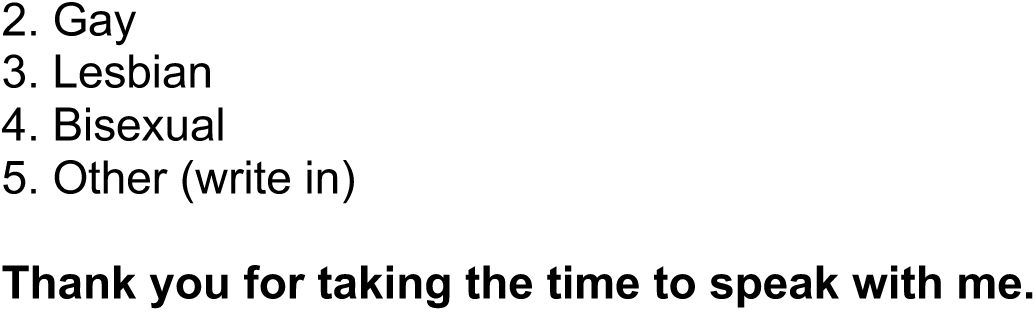
Qualitative Discussion Guide.

Largely due to COVID-related limitations, the semi-structured interviews were performed over the phone in English. Interviews were conducted until thematic saturation was reached, meaning no new findings were identified from participants. The interviews were audio recorded and transcribed. A codebook was developed combining emic and etic terms to capture both the grounded perspectives of participants and the priorities of the research team. Dedoose, a secure, HIPAA-compliant application used to analyze qualitative research based on grounded theory, was utilized to code and conduct content analysis, identifying common themes and patterns. The same 3 team members who conducted the interviews independently coded the transcriptions.

The study protocol and consent form were approved by the University of Rochester Research Subjects Review Board (RSRB) Office for Human Subject Protection (STUDY00006604).

## Results

87 patients were contacted, and 17 participants were included in the study. The duration of the interviews varied from 15-45 minutes. The demographic characteristics of the 17 Asian American participants are displayed in Table 1. The majority of participants were between 30-40 years old. All were assigned female sex at birth and identified their gender as female. Most participants reported they were heterosexual or straight. Participants identified affiliations with a variety of Asian American ethnicities, with “Chinese” being the most common.

**Table 1.** Demographic characteristics of Asian American participants.

| **Demographic** |  | **N** |
| --- | --- | --- |
| **Age (years)** | 18-29 | 0 |
|  | 30-39 | 9 |
|  | 40-49 | 5 |
|  | ≥50 | 3 |
| **Sex** | Female | 17 |
| **Gender Identity** | Female | 17 |
| **Sexual Identity** | Heterosexual or straight | 15 |
|  | Gay | 0 |
|  | Lesbian | 0 |
|  | Bisexual | 2 |
|  | Other | 0 |
| **Ethnicity** | Chinese | 4 |
|  | Indian | 3 |
|  | Filipino | 2 |
|  | Vietnamese | 1 |
|  | Korean | 1 |
|  | Japanese | 1 |
|  | Pakistani | 0 |
|  | Thai | 0 |
|  | Cambodian | 0 |
|  | Hmong | 0 |
|  | Laotian | 1 |
|  | Bangladeshi | 0 |
|  | Nepalese | 1 |
|  | Burmese | 1 |
|  | Indonesian | 0 |
|  | Sri Lankan | 0 |
|  | Malaysian | 0 |
|  | Mongolian | 0 |
|  | Other | 2 |

Figure 2 displays the 6 significant themes were extracted from the interviews. The themes were not predetermined and derived directly from the data.

**Figure 2.**
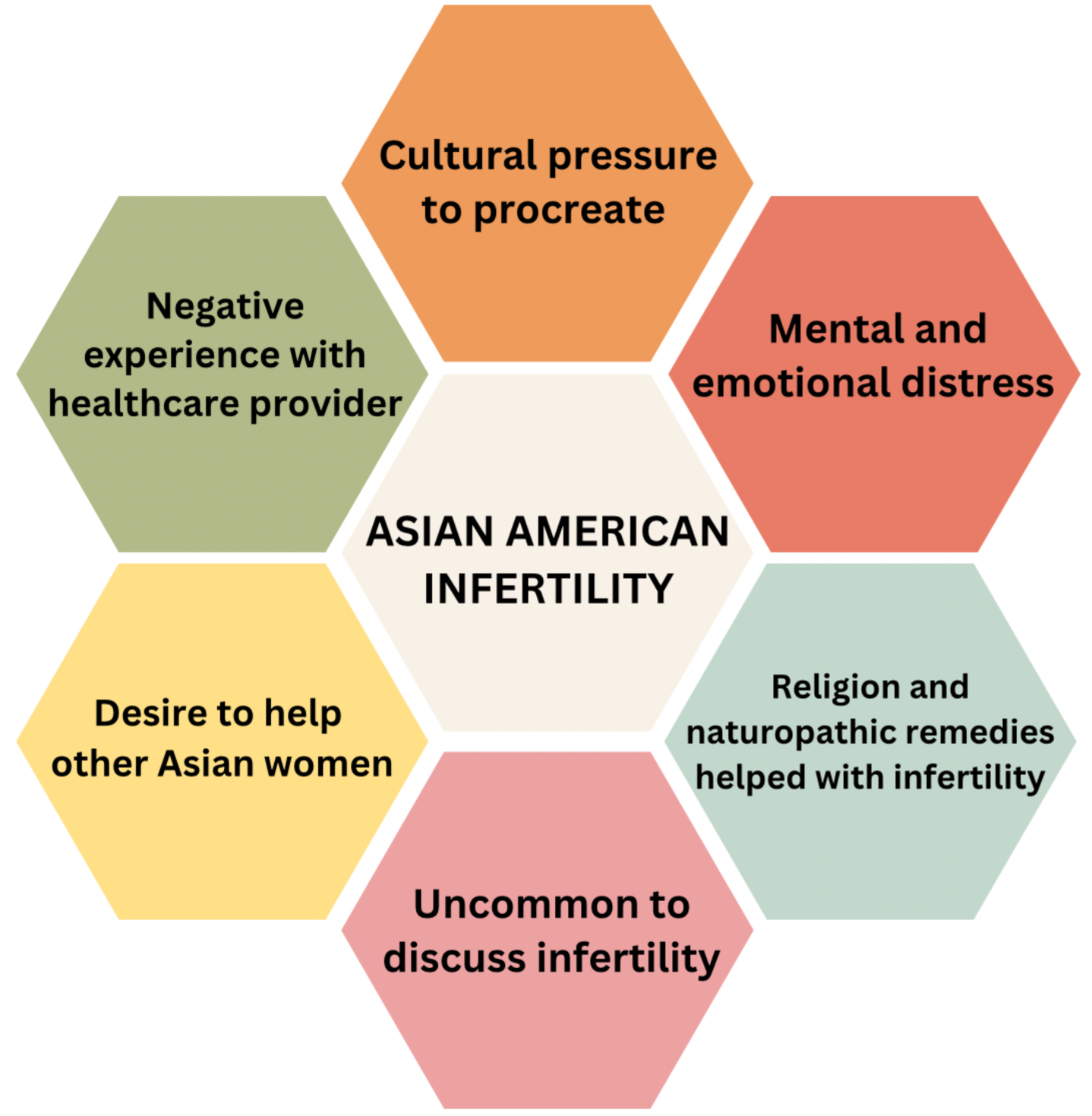
Significant themes from qualitative interviews.

### Theme 1: Cultural pressure to procreate

Participants voiced the importance of having children, emphasizing the cultural expectation to procreate. Women persistently reported feeling a personal responsibility to bear children.

“I feel like Asian people are supposed to have like the women are supposed to have kids…like part of what you’re supposed to do is have kids like that’s a big deal, like you’re supposed to make baby like, take care and have children so they can have their own kids.” – Korean female in her 40s

“…if you don’t get pregnant, if you don’t have children, they don’t say anything but you know that they’re aware and look down on you.” – Chinese female in her 40s

“So in our culture is like, different like where we are from…we have some kind of like different thinking like all traditional thinking. So like when you don’t have children like they think that you are – that is not good luck in their house.” – Nepalese female in her 30s

Participants also reported that it would be culturally appropriate for a man to leave his wife if she is unable to reproduce, resulting in increased pressure to conceive.

“…if they [husbands] don’t have children, what do they do then? They won’t keep the marriage. They won’t keep the promises…if you don’t have a child, you will get a divorce.” – Chinese female in her 50s

“Well, so for those who can’t get pregnant, they will be so worried of security. A few years after getting married, they can’t get pregnant or they discovered that they have any problem, there is the chance that the husband will get another wife. He has the right-like we accept that, you know.” – Pakistani female in her 40s

### Theme 2: Mental and emotional distress

Every participant reported that infertility caused immense mental and emotional distress. They reported feelings of disappointment and grief were verbalized multiple times.

“The hopelessness. There is no hope to have your own baby. By the time you see the doctor, every month, I felt hopeless.” – Chinese female in her 30s

“Knowing that I can’t carry a child makes me feel, um for awhile, makes me feel worthless… depression is mainly like why, why me and what did I do wrong or how did this happen to me, you know?” – Vietnamese female in her 30s

The high cost of infertility evaluation and treatment caused a particularly large amount of distress.

“And financially, I think, you know, my first child, my husband and I calculated we had spent one hundred and seven thousand dollars before she was born, and that was a lot of money for just fertility treatments.” – Indian female in her 30s

“So we actually we looked online when we started talking about donor eggs and all that. I mean, it’s like, it’s like-it was $14,000-15,000 for like six eggs. I mean, the cost was just insane and there’s certain things about insurance…there are certain things insurance doesn’t cover, so it wouldn’t cover donor eggs…and that, you know, I think the cost of that was really challenging and stressful. You have to do it within a certain period of time and it was just expensive.” – Korean female in her 40s

One patient even sought care outside of the United States due to the high cost of IVF.

“Money is a cause of concern…I had a body, my body checked and I had a surgery in U.S. and then we didn’t continue the IVF program. We decided I would take it, the IVF program, in Taiwan.” – Taiwanese female in her 40s

### Theme 3: Uncommon to discuss infertility

Participants noted that it is not common in their cultures to talk about infertility or that infertility was not commonly discussed in the past. However, they did mention that infertility is becoming more openly discussed presently, though most still did not tell family or friends about their own infertility experience.

“I think infertility in Japanese culture, anyways, is like not something that we really speak about.” – Japanese female in her 30s

“Just the thought of just nobody talks about it like it’s always so easy, it should be easy to have a baby… like we don’t talk about it even in my in, I guess, culture…I didn’t really say much about it to my family and stuff like that.” – Laotian female in her 40s

“It used to be really uncommon, but now I think more and more people are opening up to it, especially our generation. I bet in my parents’ generation it wasn’t at all common to talk about it…and in my generation, I want to say there’s also, you know, more acceptance and more empathy towards whoever is going through it…my husband and I were the only ones who knew about me.” – Indian female in her 30s

### Theme 4: Negative experience with healthcare provider

Participants reported being unable to build a strong, trusting relationship with their providers. One specific area of frustration was the lack of individualized care.

“I feel like I was just a number coming in to get it done. So I didn’t feel the love of like caring and personal service for me…so that wasn’t making me feel comfortable, you know? And um, when I asked for information, yes information is provided, but it doesn’t all pertain to me. She just gave a general of what the standard will be. Well, you know, it’s just not personalized to me.” – Vietnamese female in her 30s

“I thought um there could have been better guidance… I don’t know how much, you know, a holistic lifestyle helps…every time I would ask, the answer would be brushed off…maybe this is not a strong correlation, but these things don’t hurt to answer my questions.” – Indian female in her 30s

“I feel, yeah, it’s very hard for doctor to tell the patient that the pregnancy was the absolutely that their pregnancy cannot shouldn’t keep going…it’s very hard for patients to hear that, especially on the table when they are on the bed for ultrasound…I think maybe at that time should work on the communication better…I think they should spend more time for patients understand the benefits and disadvantages.” – Chinese female in her 40s

### Theme 5: Religion and naturopathic remedies helped with infertility

Religion played a large role in helping participants adjust to their infertility journeys even if they didn’t consider themselves to be very religious.

“We are not that much religious…but I believe to Jesus, so I really prayed to God.” –Indian female in her 40s

“I don’t have a very strong connection to my spiritual community right now, but I remember during my treatment, I like went to weekday mass, which I rarely ever do, but it was a source of comfort…there are times when I turn to prayer when I was really struggling.” – Fillipino female in her 30s

“Right when we started this journey, we put our absolute faith in science… but there’s probably still something beyond science or beyond control. It’s something that cannot be explained, and it almost still feels like God’s will…I don’t know what else God does or doesn’t do, but life and death, at least, are definitely you know his domain.” – Indian female in her 30s

Respondents also turned to naturopathic remedies, including acupuncture and herbal medicine, to aid in their infertility treatment.

“I actually like some of the medication I bought from China and India. I thought that helps me too at that point. To keep my immuno high.” – Nepalese female in her 30s

“I did exercise…I went to some yoga classes… I tried acupuncture.” – Vietnamese female in her 30s

### Theme 6: Desire to help other Asian women

Most participants expressed desire to help other Asian women who are also struggling with infertility. Many were interested in joining support groups for Asian Americans facing infertility.

“I tried to help the future people who might feel the same way as I do to kind of, you know, like, it’s OK. It’s OK to talk about, it’s OK to feel this way, you know, you’re not alone kind of thing” – Japanese female in her 30s

“[it’s hard] not having somebody who’s going through the same thing, like another woman who’s going through the same thing to talk to…so I think it would have really it would have helped my journey if I just had a couple of girls who were going through the same thing as me.” – Indian female in her 30s

“I know there are so many people are having the same issue like us, you know, so if there’s anything we could do to help with the research, you know, that kind-that kind of thing. We would love to.” – Chinese female in her 30s

## Discussion

This study highlighted the complexity and difficulty of the Asian American infertility experience. Parenthood is a strong expectation for people of Asian descent and results in a mental and emotional burden that appears to disproportionately impact women. In East Asia, matrimony and childbearing are commonly viewed as paramount life events, and family building allows young adults to attain moral significance and status.[15] One of the proposed reasons having children is so important is because Asian people may see children as ‘social security’ with the idea that children will take care of their parents when they become older.[16, 17] As the women in our study repeatedly echoed, the cultural belief that Asian women are responsible to procreate places enormous pressure on women diagnosed with infertility. To add to the psychologic distress, participants in our study expressed the fear that their partners would leave them if they were infertile. Yet women had few modes of communication available with healthcare professionals, family, or friends through which to express these pressures. In a study of women diagnosed with infertility in Tianjin, China, women similarly reported that childbearing was essential to preventing marital failure and/or rejuvenating an unsatisfactory marital relationship.[18] Moreover, in a meta-analysis of 25 studies investigating the prevalence of intimate partner violence (IPV) among women living in Africa and West, South and East Asia, with infertility, infertile women are more likely to experience IPV, including both psychologic and physical violence, when compared to women without infertility.[19]

Due to the sense of shame associated with infertility, people of Asian descent are unlikely to discuss their fertility journeys. In general, across most cultures, health concerns regarding reproduction and fertility are sensitive and considered taboo to discuss.[20] It has also been suggested that in this population discussion of any general health concerns is seen as a weakness and makes them appear fragile.[21] However, while the majority of participants in our study did not tell their family and friends about their infertility, they commented on their belief that infertility is more commonly discussed at present than in the past. This change may be due to the fact that medical professionals and organizations have normalized infertility experiences as medical phenomena. In 2009, the World Health Organization (WHO) defined infertility as “a disease of the reproductive system” for the first time.[22] The increased awareness of infertility as a prevalent medical condition and public health issue may encourage patients to speak more freely about their reproductive challenges.

As mentioned previously, Asian Americans are less likely to report strong relationships with their healthcare providers. When reflecting on their infertility experiences, many of our participants reported frustration with their providers’ communication styles, disregarding their questions and rushing through counseling. This disregard appears to be a common theme in the literature as multiple studies have reported that Asian patients are more likely to be dissatisfied with their medical care compared to their white counterparts.[23, 24] These feelings may result in lack of or delayed infertility evaluation and treatment, which has been seen in other aspects of women’s healthcare. For example, Vietnamese American women are five times more likely to have cervical cancer compared to white women, but only half of Vietnamese women aged 18 years and older report ever having a Pap smear.[25]

Due to their longstanding history of practicing medicine rooted in Asian societies, Asian Americans are more likely to use complementary and alternative medicine (CAM) compared to other race/ethnicities.[26, 27] The Asian American women in our study reported utilizing a wide range of complementary treatments, including herbal medications, acupuncture, and specific exercises, including tai chi and yoga. Although CAM such as acupuncture may not result in increased live birth rates when treating infertility, they may provide other benefits such as reducing stress and anxiety.[28–29] Healthcare providers can work to discuss and integrate culturally meaningful naturopathic remedies into infertility treatment plans in an effort to alleviate cultural barriers and provide individually centered care. This integration may aid in building stronger physician-patient relationships.

The Asian American women in our study repeatedly expressed their desire to help other Asian women also experiencing infertility. In-person support groups have been found to be an effective intervention to reduce the stress associated with infertility treatment.[30] However, utilization of social media and online forums should also be considered to reduce logistical barriers. Participants of online infertility support groups have reported increased access to information, reduced sense of isolation and increased empowerment.[31]

Our study is not without limitations. One limitation of the study is the small sample size with only 17 patients interviewed. While thematic saturation was reached, the sample size still limits the generalizability of the study results. It has been documented that Asian American patients are less likely to participate in infertility research for a variety of reasons, including the belief that research is associated with compromised clinical care.[6, 32] Our study was only conducted in English and thus excludes patients who cannot speak this language. Utilizing interpreters may help to increase Asian American involvement in future studies. One of the largest limitations of our study is the use of the monolithic term “Asian” when collecting demographic data. According to the U.S. Census Bureau, when a person checks “Asian” in the race category, they are “a person having origins in any of the original people of the Far East, Southeast Asia, or the Indian subcontinent, including, for example, Cambodia, China, India, Japan, Korea, Malaysia, Pakistan, the Philippine Islands, Thailand, and Vietnam”.[3] This encompasses a very broad range of ethnicities, each of which may have their own infertility patterns and trends. Future studies could benefit from investigating the trends that differentiate racial and ethnic categories within the term “Asian” to prevent overgeneralization of findings and better individualize patient care.

Our study provides more insight to the unique barriers Asian Americans face when diagnosed with infertility and seeking fertility care. Healthcare providers may consider addressing the cultural pressure Asian American patients face and its impact on their mental/emotional health. By increasing research of naturopathic remedies and discussing the importance of religion with patients, medical professionals can strengthen the physician-patient relationship. Helping connect Asian American women with one another can also build social support.

## Conclusion

Asian Americans commonly experienced culturally-determined stigma and distress around reproduction, with fewer perceived opportunities to communicate effectively about their infertility experience with care providers, family or friends. They expressed a desire to support other women experiencing infertility and explored religion and traditional therapies to address their fertility issues.

## Data Availability

All relevant data are within the manuscript and its Supporting Information files.

## Notes

**Funding statement:** There was no funding for this research study.

### Competing Interest Statement

The authors have declared no competing interest.

### Funding Statement

The author(s) received no specific funding for this work.

